# How does public knowledge, attitudes and behaviours correlate in relation to COVID-19? A community-based cross-sectional study in Nepal

**DOI:** 10.1101/2020.07.30.20165266

**Authors:** Hridaya Raj Devkota, Tula Ram Sijali, Ramji Bogati, Andrew Clarke, Pratik Adhikary, Rajendra Karkee

## Abstract

**Background:** The COVID-19 pandemic has created a global health emergency requiring an effective public health response including citizen’s roles in preventing spread and controlling the pandemic. Little is known about public knowledge, beliefs and behaviors in-relation to the pandemic in Nepal. This study aims to assess knowledge, attitude and practices (KAP) towards COVID-19 among the general public and to identify associated factors.

**Methods:** A cross-sectional survey was conducted between May - June 2020 with a sample of 645, recruited from 26 hospitals across Nepal. We conducted telephone interviews using a semi-structured questionnaire related to KAP regarding COVID-19. T-test and one-way ANOVA was conducted to determine group differences for socio-demographic variables. Linear regression and correlational analysis were performed to identify associated factors and measure strength and direction of relationships.

**Results:** Overall mean scores for knowledge, attitude and practice were 11.6 (SD 4.5), 2.7 (SD 1.8), and 9.9 (SD 1.93) respectively but differed by socio-demographic characteristics. Positive but weak linear correlations were observed between knowledge-practice (r=0.19, p<0.01) and attitude-practice (r=0.08, p<0.05). The relationship between knowledge and education was fairly strong (r = 0.34, p< 0.01). Province, place of residence, ecological area, age, gender and caste/ethnicity were also significantly associated with KAP score of participants.

**Conclusion:** The study found varying degrees of correlation between Knowledge, Attitude and Practice that may increase as the pandemic evolves in Nepal. Knowledge and level of education had positive associations with attitude and adherence of precautionary measures. The findings suggest a need for targeted community awareness interventions for the most vulnerable populations, men, those with no school education, the elderly and people living in rural areas.

## Background

The World Health Organization (WHO) declared COVID-19 to be a public health emergency on 30th January 2020, after a month of the Corona virus outbreak in Wuhan, China (1). Nepal detected the first case of Corona virus infection on 23rd January that surged to 11,700 affecting all 77 districts with a total 28 reported deaths by the end of June 2020 (2–4). Measures including a country-wide lock down have been adopted to prevent transmission, however the disease continues to spread.

Prevention and case-management during pandemics requires public support together with government action. The effectiveness of actions and control measures depends on the extent to which people change their behavior. The health belief theory explains that a person is likely to take health actions if the individual believes that s/he is susceptible to the disease or would have serious effect upon him if contracted. Further, if a person is aware about certain actions that can be taken and believe that these actions may reduce his likelihood of contracting or reduce the severity of disease (5). Literature informs that individual beliefs and perceptions play an important role in subsequent behavioral change (6).

Studies conducted during the SARS outbreak in 2003 found individual beliefs and perceptions to be an important factor in subsequent behavior change (7). Moreover, that the higher the perceived effectiveness of measures, the higher the chances of action being undertaken. Likewise, higher perceived threats of the disease lead to higher rates of behavioral change (8).

Having a well-informed public about the COVID-19 virus, it’s causes and mode of transmission, could be one of the best strategies to prevent and slow transmission. However, until recently there was limited information and scientific knowledge about the virus. Scientific understanding about mutation rate, transmission, disease symptoms and severity, herd immunity and risk groups is still emerging and this uncertainty creates a challenge for reliably informing the public, resulting in confusion about the best practices for health protection and negative impacts on mental health (9). Under such conditions, designing effective and contextually appropriate interventions to support risk reduction and behavior change is demanding.

There is an urgent need to understand public knowledge about COVID-19, their beliefs and behaviors, in order to produce information that facilitates effective public health responses. This study aimed to assess the knowledge, attitude and practice towards COVID-19 among the Nepali population and identify any relationships between KAP scores and demographic factors.

## Methodology

### Study design and population

A cross-sectional survey among attendants to fever clinics in hospitals was conducted between May 17 and June 9, 2020.

### Participants’ recruitment procedure

A multi-stage sampling method was used for recruitment of participants. The study covered all seven provinces, with participants drawn from 26 health facilities, from 23 out of 77 districts covering both ecological zones – hills and Terai. A sampling frame was developed collecting the names of those who attended fever clinics between April 25 to May 16, 2020 in the selected health facilities. Out of 1,285 fever clinic attendants, 687 met the eligibility criteria for the study and were included in the study.

Individuals aged 18 and above, who visited hospital suspecting or having COVID-19 symptoms were the inclusion criteria. The response rate was 84%.

### Survey instrument and data collection procedure

A semi-structured questionnaire seeking socio-demographic information and knowledge, attitudes and practices (KAP) regarding COVID-19 was developed and administered. The socio-demographic information included participant’s age, gender, education, occupation, caste and ethnicity, religion and marital status. The living area (province), ecological zone and place of residence (urban or rural) were included. The second section of the questionnaire consisted of knowledge about COVID-19, attitudes and practices regarding COVID-19. Based on the published literature, WHO and local government’s information and guidelines (10–12) for COVID-19, 27-items of knowledge, 3-items of attitude and 4-items of practice related questions were adapted.

The standardized questionnaire was set up on tablet computers and mobile phones with KoBo Collect software and administered in Nepali through telephone interviews by trained data collectors. The questionnaire was first developed in English, translated into Nepali by three bilingual Nepalese and field-tested for acceptability and comprehension among the population in which it was to be used. On average, administration of the questionnaire took 24 minutes.

The research obtained ethical approval from the Nepal Health Research Council (NHRC) - ERB Protocol Registration No. 317/2020P. Before interviews, verbal informed consent was taken from all participants.

### Measures

The KAP indicators were created by questionnaire items to derive scores. The knowledge questionnaire consisted of 9 items about COVID-19 symptoms, 2 items about risk, and 8 items each about transmission and prevention. Knowledge in those questions was spontaneously cited and presented as an additive score. All the questionnaire items were equally weighted, dichotomized, and score was created using the sum with the maximum scores of 27. The attitude score was developed using a 3-item questionnaire about the individual beliefs on remaining safe from COVID-19, beliefs on easy availability of healthcare services, and belief on government’s ability to control the current pandemic. Similarly, the practice questionnaire included questions about social distancing, use of masks, hand washing and use of hand sanitizer. The two practice questions had two rating scales while all the other attitude and practice items consisted of four rating scales with the score weight ranging 0 – 3 making a maximum total score of 6 and 12 for attitude and practice respectively. In this study it is interpreted that the higher the attitude score, the higher the pessimistic attitude or perceived risk. Chronbach’s Alpha coefficient of the knowledge and attitude questionnaires were calculated 0.78 and 0.71 respectively indicating acceptable internal consistency (13).

### Statistical Analysis

This study analyzed the data using SPSS (version 23.0 for Windows). We used the descriptive and inferential statistics. The categorical variables were summarized using frequency and percentage, and the continuous variables using mean and standard deviation (SD). Independent sample t-test and one-way analysis of variance (ANOVA) were conducted to determine the differences between groups for selected socio-demographic variables, while bivariate correlation analysis was performed to measure the strength and direction of relationship. The relationships between knowledge, attitudes and practice scores were examined using bivariate correlational analyses and multivariate linear regression models. We conducted linear regression analysis using knowledge, attitude and the demographic variables as independent variables and practice score as the outcome variable to identify factors associated with practice.

## Results

### Characteristics of study participants

Out of 687 people approached for interview, 6% refused and 645 participants were interviewed. 27 interviews were excluded from analysis due to incomplete information and a total of 618 were included in the analysis. The highest proportion of participants (17.8%) were from Karnali (Province 6) and the lowest from Province 5. The majority of study participants (79%) lived in urban area, while 63.6% in the hills. The average age of the participants was 35 years ranging 18 – 85 (SD = 14.25). More than one-third (37%) were women, majority of the participants (55%) reported having their secondary level education and 16% with higher education. Nearly 4 in 10 reported their occupation as labor and 16.8% having foreign employment. Just under 43% participants reported their caste group as Brahmin/Chhetri and over 17% as Dalits, and 77% were married. (Table 2)

### Knowledge of COVID-19

The study found COVID-19 knowledge mean score at 11.6 (SD 4.5; range 0 – 27) suggesting an overall knowledge score rate at 43%. Knowledge score significantly differed among the participants by province, residence area, age, gender, education, occupation and marital status. Province 5 respondents had highest rate of knowledge score (mean 12.8; SD 5.6) followed by Gandaki (mean: 12.4, SD 4.6) and Province 2 (mean: 12.4, SD 5.0), while Karnali people had the least (mean: 10.4, SD 4.4). Urban dwellers scored higher (mean 11.9, SD 4.5) than rural (mean 10.4, SD 4.3). Similarly, males scored higher (mean 11.9, SD 4.4) than females (mean 11.1, SD 4.7). Among the age groups, participants 25 – 34 years scored the highest (mean 12.2, SD 4.4) followed by 35 – 44 age groups (mean 11.8, SD 4.3), and those aged over 55 years scored the lowest (mean 9.7, SD 4.7). Participants with higher education scored higher (mean 14.2, SD4.2) than those with no school education (mean 8.9, SD 3.9). Participants who reported their main occupation as service had highest knowledge scores (mean 13.5, SD 4.3), followed by the business or self-employed group (mean 12.5, SD 3.8), while the farmers scored the lowest (mean 10.4, SD 4.8).

Ever married participants scored highest (mean 12.5, SD 4.4), while single (widowed or divorced) scored the lowest (mean 7.8, SD 4.4). (Table 1)

**Table 1:** Knowledge score of COVID-19 by demographic characteristics (n = 618)

| Variables | Frequency (%) | Mean | SD | t/F - Ratio | P-value |
| --- | --- | --- | --- | --- | --- |
| Province |  |  |  |  |  |
| Karnali_6 | 110 (17.8) | 10.4 | 4.4 | 3.72 | 0.001 |
| Gandaki_4 | 107 (17.3) | 12.4 | 4.6 |  |  |
| Bagmati_3 | 96 (15.5) | 10.7 | 3.8 |  |  |
| Province_1 | 92 (14.9) | 11.8 | 4.3 |  |  |
| Sudurpaschim_7 | 80 (12.9) | 11.2 | 3.8 |  |  |
| Province_2 | 75 (12.1) | 12.4 | 5.0 |  |  |
| Province_5 | 58 (9.4) | 12.8 | 5.6 |  |  |
| Ecological region |  |  |  |  |  |
| Hill Districts | 393 (63.6) | 11.4 | 4.5 | -1.53 | 0.127 |
| Terai Districts | 225 (36.4) | 11.9 | 4.5 |  |  |
| Place of Residence |  |  |  |  |  |
| Urban Municipality | 488 (79.0) | 11.9 | 4.5 | 3.39 | 0.001 |
| Rural Municipality | 130 (21.0) | 10.4 | 4.3 |  |  |
| Age |  |  |  |  |  |
| 18 - 24 | 151 (24.4) | 11.7 | 4.5 | 5.28 | 0.000 |
| 25 - 34 | 221 (35.8) | 12.2 | 4.4 |  |  |
| 35 - 44 | 123 (19.9) | 11.8 | 4.3 |  |  |
| 45 - 54 | 48 (7.8) | 10.6 | 4.5 |  |  |
| 55 and above | 75 (12.1) | 9.7 | 4.7 |  |  |
| Gender |  |  |  |  |  |
| Male | 390 (63.1) | 11.9 | 4.4 | 2.08 | 0.038 |
| Female | 228 (36.9) | 11.1 | 4.7 |  |  |
| Caste & Ethnicity |  |  |  |  |  |
| Brahmin/Chhetri | 264 (42.7) | 11.9 | 4.2 | 1.89 | 0.129 |
| Jana Jaati | 160 (25.9) | 11.7 | 4.6 |  |  |
| Dalit | 110 (17.8) | 10.9 | 5.1 |  |  |
| Madhesi/Muslims & others | 84 (13.6) | 11.0 | 4.4 |  |  |
| Education |  |  |  |  |  |
| No formal education | 98 (15.9) | 8.9 | 3.9 | 27.94 | 0.000 |
| Primary (1 - 5) | 84 (13.6) | 10.3 | 4.5 |  |  |
| Secondary (6 - 12) | 340 (55.0) | 11.9 | 4.3 |  |  |
| Higher education | 96 (15.5) | 14.2 | 4.2 |  |  |
| Occupation |  |  |  |  |  |
| Labor and others | 240 (38.8) | 11.3 | 4.4 | 9.08 | 0.000 |
| Service | 106 (17.2) | 13.5 | 4.3 |  |  |
| Foreign Employment | 104 (16.8) | 10.7 | 4.6 |  |  |
| Farming | 94 (15.2) | 10.4 | 4.8 |  |  |
| Business & self-employment | 74 (12.0) | 12.5 | 3.8 |  |  |
| Marital status |  |  |  |  |  |
| Married | 475 (76.9) | 11.4 | 4.5 | 8.82 | 0.000 |
| Ever Married | 127 (20.6) | 12.5 | 4.4 |  |  |
| Single (Widow, Divorced) | 16 (2.6) | 7.8 | 4.4 |  |  |

**Table 2:** Attitude towards COVID-19 score by demographic characteristics (n = 618)

| Variables | Frequency (%) | Mean | SD | t/F - Ratio | P-value |
| --- | --- | --- | --- | --- | --- |
| **Province** |  |  |  |  |  |
| Karnali_6 | 110 (17.8) | 3.7 | 2.0 | 11.76 | 0.000 |
| Gandaki_4 | 107 (17.3) | 3.0 | 1.5 |  |  |
| Bagmati_3 | 96 (15.5) | 3.0 | 1.6 |  |  |
| Province_1 | 92 (14.9) | 2.0 | 1.5 |  |  |
| Sudurpaschim_7 | 80 (12.9) | 2.4 | 1.9 |  |  |
| Province_2 | 75 (12.1) | 2.0 | 1.8 |  |  |
| Province_5 | 58 (9.4) | 2.8 | 1.7 |  |  |
| **Ecological region** |  |  |  |  |  |
| Hill Districts | 393 (63.6) | 3.1 | 1.8 | 6.53 | 0.000 |
| Terai Districts | 225 (36.4) | 2.1 | 1.7 |  |  |
| **Place of Residence** |  |  |  |  |  |
| Urban Municipality | 488 (79.0) | 2.7 | 1.8 | -1.97 | 0.049 |
| Rural Municipality | 130 (21.0) | 3.0 | 1.9 |  |  |
| **Age** |  |  |  |  |  |
| 18 - 24 | 151 (24.4) | 2.8 | 2.0 | 3.68 | 0.006 |
| 25 - 34 | 221 (35.8) | 3.0 | 1.8 |  |  |
| 35 - 44 | 123 (19.9) | 2.6 | 1.7 |  |  |
| 45 - 54 | 48 (7.8) | 2.3 | 1.6 |  |  |
| 55 and above | 75 (12.1) | 2.3 | 1.6 |  |  |
| **Gender** |  |  |  |  |  |
| Male | 390 (63.1) | 2.8 | 1.9 | 0.33 | 0.740 |
| Female | 228 (36.9) | 2.7 | 1.7 |  |  |
| **Caste & Ethnicity** |  |  |  |  |  |
| Brahmin/Chhetri | 264 (42.7) | 2.8 | 1.7 | 1.75 | 0.155 |
| Jana Jaati | 160 (25.9) | 2.7 | 1.8 |  |  |
| Dalit | 110 (17.8) | 2.9 | 2.0 |  |  |
| Madhesi/Muslims & others | 84 (13.6) | 2.4 | 1.9 |  |  |
| **Education** |  |  |  |  |  |
| No formal education | 98 (15.9) | 2.4 | 1.8 | 6.74 | 0.000 |
| Primary (1 - 5) | 84 (13.6) | 2.4 | 1.9 |  |  |
| Secondary (6 - 12) | 340 (55.0) | 2.7 | 1.8 |  |  |
| Higher education | 96 (15.5) | 3.4 | 1.5 |  |  |
| **Occupation** |  |  |  |  |  |
| Labor and others | 240 (38.8) | 2.4 | 1.7 | 8.02 | 0.000 |
| Service | 106 (17.2) | 3.6 | 1.7 |  |  |
| Foreign Employment | 104 (16.8) | 2.7 | 1.9 |  |  |
| Farming | 94 (15.2) | 2.5 | 1.8 |  |  |
| Business & self-employment | 74 (12.0) | 2.9 | 1.7 |  |  |
| **Marital status** |  |  |  |  |  |
| Married | 475 (76.9) | 2.7 | 1.8 | 3.73 | .025 |
| Ever Married | 127 (20.6) | 3.1 | 1.8 |  |  |
| Single (Widow, Divorced) | 16 (2.6) | 2.1 | 1.9 |  |  |

### Attitude towards COVID-19

The overall attitude mean score among the study participants was 2.7 (SD 1.8, range 0 – 6). In this study it is interpreted that the higher the attitude score the higher the perceived risk. Attitude scores differed across groups (Table 2). Residents in Karnali had the highest attitude score (mean 3.7, SD 2.0), while Province 1 and 2 scored the lowest (mean 2.0, and SD 1.5 and 1.8 respectively). Participants aged between 25 – 34 had the highest attitude score (mean 3.0, SD 1.8), while those 45 – 54 and 55 + scored the lowest (mean 2.3, SD 1.6). Similarly, participants having higher education had higher score (mean 3.4, SD 1.5) than those with no school education. Respondents employed in the service sectors had higher attitude score (mean 3.6, SD 1.7) while the labor group had the lowest (mean 2.4, SD 1.7). (Table 2)

### Practice towards COVID-19

The participant’s overall mean practice score was 9.9 (SD 1.93, range 3 – 12). However, the score differed between groups (Table 3). Residents in Gandaki had the highest score (mean 10.8, SD 1.6) and Karnali reported the lowest (mean 9.4, SD 1.7). Urban respondents scored slightly more (mean 10.0, SD 1.9) than rural (mean 9.5, SD 1.9), and female (mean 10.2, SD 1.9) more than male (mean 9.7, SD 2.0). Among caste and ethnic groups, Bramhin/Chhetri had the highest practice score (mean 10.1, SD 1.9) and the lowest were found among Dalit respondents (mean 9.5, SD 1.9). Those with higher education level had the highest score (mean 10.7, SD 1.6), and the lowest with no school education or primary level education (mean 9.2, SD 2.0). Respondents working in service occupations had the highest practice score (mean 10.5, SD 1.7) and again, farmers scored the lowest (mean 9.2, SD 1.9). (Table 3)

**Table 3:** Practice towards COVID-19 score by demographic characteristics (n = 618)

| Variables | Frequency (%) | Mean | SD | t/F - Ratio | P-value |
| --- | --- | --- | --- | --- | --- |
| **Province** |  |  |  |  |  |
| Karnali_6 | 110 (17.8) | 9.4 | 1.7 | 7.93 | 0.000 |
| Gandaki_4 | 107 (17.3) | 10.8 | 1.6 |  |  |
| Bagmati_3 | 96 (15.5) | 10.1 | 1.9 |  |  |
| Province_1 | 92 (14.9) | 9.5 | 2.2 |  |  |
| Sudurpaschim_7 | 80 (12.9) | 9.3 | 1.9 |  |  |
| Province_2 | 75 (12.1) | 9.9 | 1.9 |  |  |
| Province_5 | 58 (9.4) | 9.9 | 2.0 |  |  |
| **Ecological region** |  |  |  |  |  |
| Hill Districts | 393 (63.6) | 9.8 | 2.0 | -0.92 | 0.357 |
| Terai Districts | 225 (36.4) | 10.0 | 1.9 |  |  |
| **Place of Residence** |  |  |  |  |  |
| Urban Municipality | 488 (79.0) | 10.0 | 1.9 | 2.51 | 0.012 |
| Rural Municipality | 130 (21.0) | 9.5 | 1.9 |  |  |
| **Age** |  |  |  |  |  |
| 18 - 24 | 151 (24.4) | 10.0 | 1.9 | 0.45 | 0.773 |
| 25 - 34 | 221 (35.8) | 9.8 | 1.9 |  |  |
| 35 - 44 | 123 (19.9) | 9.8 | 1.9 |  |  |
| 45 - 54 | 48 (7.8) | 9.6 | 2.0 |  |  |
| 55 and above | 75 (12.1) | 9.9 | 1.9 |  |  |
| **Gender** |  |  |  |  |  |
| Male | 390 (63.1) | 9.7 | 2.0 | -2.99 | 0.003 |
| Female | 228 (36.9) | 10.2 | 1.9 |  |  |
| **Caste & Ethnicity** |  |  |  |  |  |
| Brahmin/Chhetri | 264 (42.7) | 10.1 | 1.9 | 2.61 | 0.050 |
| Jana Jaati | 160 (25.9) | 9.7 | 2.0 |  |  |
| Dalit | 110 (17.8) | 9.5 | 1.9 |  |  |
| Madhesi/Muslims & others | 84 (13.6) | 10.0 | 2.1 |  |  |
| **Education** |  |  |  |  |  |
| No formal education | 98 (15.9) | 9.2 | 2.0 | 14.48 | 0.000 |
| Primary (1 - 5) | 84 (13.6) | 9.2 | 1.9 |  |  |
| Secondary (6 - 12) | 340 (55.0) | 10.0 | 1.9 |  |  |
| Higher education | 96 (15.5) | 10.7 | 1.6 |  |  |
| **Occupation** |  |  |  |  |  |
| Labor and others | 240 (38.8) | 10.0 | 2.0 | 11.14 | 0.000 |
| Service | 106 (17.2) | 10.5 | 1.7 |  |  |
| Foreign Employment | 104 (16.8) | 9.2 | 1.9 |  |  |
| Farming | 94 (15.2) | 9.2 | 1.9 |  |  |
| Business & self-employment | 74 (12.0) | 10.4 | 1.7 |  |  |
| **Marital status** |  |  |  |  |  |
| Married | 475 (76.9) | 9.8 | 2.0 | 2.55 | .079 |
| Ever Married | 127 (20.6) | 10.2 | 1.8 |  |  |
| Single (Widow, Divorced) | 16 (2.6) | 9.8 | 1.8 |  |  |

### Correlation between knowledge, attitude, practice and demographic characteristics

This study interpreted the correlations criteria with r value 0 – 0.25 = a weak correlation, 0.25 – 0.5 = fair correlation, 0.5 – 0.75 = moderate correlation, and >0.75 = strong correlation (14). Table 4 shows the correlation between KAP scores and their relation with demographic characteristics. The analysis of knowledge-practice (r = 0.19, p < 0.01) and attitude-practice (r = 0.08, p < 0.05) both showed significant positive linear correlations, however they were weak. (Table 4)

**Table 4:** Correlation matrix among interest variables (n=618)

|  | Pearson/Spearman's rho correlation coefficient |  |  |
| --- | --- | --- | --- |
|  | Knowledge | Attitude | Practice |
| Knowledge | - |  |  |
| Attitude | 0.04 | - |  |
| Practice | 0.19** | 0.08* | - |
| Province | 0.11** | -0.23** | -0.04 |
| Ecological area | 0.06 | -0.25** | 0.04 |
| Place of residence | -0.13** | 0.08* | -0.10* |
| Age in years | -0.11** | -0.12** | -0.03 |
| Caste and Ethnicity | -0.11** | -0.04 | -0.07 |
| Gender | -0.08* | -0.02 | 0.12** |
| Education | 0.34** | 0.16** | 0.25** |
| Primary Occupation | 0.00 | 0.06 | -0.06 |
| Marital Status | 0.04 | 0.07 | 0.07 |

\*\* Correlation is significant at the 0.01 level (2-tailed).
\* Correlation is significant at the 0.05 level (2-tailed).

The relationship between knowledge and education was fairly strong (r = 0.34, p< 0.01) but with other variables was weak. Knowledge was positively correlated with participant’s province (r = 0.11, p < 0.01) while the relationship with place of residence (r = -0.13, p < 0.01), Age (r = -0.11, p < 0.01) Caste and ethnicity (r = -0.11, p < 0.01) and gender (r = -0.08, p < 0.05) were negatively correlated. Attitude was correlated with province (r = -0.23, p< 0.01), ecological area (r = -0.25, p < 0.01), place of residence (r = - 0.08, p < 0.05), age (r = -0.12, p < 0.01) and the education level of participants (r = 0.16, p < 0.01). All of them showed negative correlation except education, however those relationships were weak. Likewise, practice score correlations were weak in relation to place of residence (r = -0.10, p <0.05), gender (r = 0.12, p <0.01) and education (r = 0.25, p < 0.01). Gender and education were positively correlated with practice, while with place of residence was negative. (Table 4)

Linear regression analysis showed that knowledge (β = -0.13, p < 0.01), province (β = -0.12, p < 0.05), ecological area (β = 0.14, p < 0.01), age (β = 0.09, p < 0.05), gender (β = 0.18, p < 0.01), and education (β=0.27, p < 0.01) significantly associated with practice score after controlling the confounders (Table 5).

**Table 5:** Result of linear regression analysis on factors associated with practices towards COVID-19

| Independent Variables | Coefficient ($\beta$) |
| --- | --- |
| Knowledge | 0.13** |
| Attitude | 0.06 |
| Province | -0.12* |
| Ecological area | 0.14** |
| Place of Residence | -0.01 |
| Age | 0.09* |
| Gender | 0.18*** |
| Caste and Ethnicity | 0.00 |
| Education | 0.27*** |
| Primary Occupation | 0.01 |
| Marital Status | 0.07 |

\* $P < 0.05$ , \*\* $P < 0.01$ , \*\*\* $P < 0.001$

## Discussion

Within a short period of time COVID-19 has had a huge impact on people’s lives. However, there has been limited research to understand public knowledge, attitudes and practices toward COVID-19 in Nepal. This study identifies correlations between KAP and population characteristics, which may inform and support planners and decision makers in policy formulation and in implementing pandemic response plans.

The study found the average score for knowledge and attitude at 43% and 45% respectively, while the average practice score was high at 82.5%. These low scores in knowledge and attitude indicate a huge gap in knowledge and attitudes relating to COVID-19. The scores differed across the background and characteristics of the sample. Participants in Karnali, living in rural areas, having no school education, who were older, female, and widowed or divorced, had the lowest knowledge scores. This contrasts with studies conducted in China and Malaysia that found older age groups and females had greater knowledge (15,16). Higher levels of knowledge about COVID-19 have also been found in studies in other Asian countries among the general public (15–17). The paradox of these findings from Nepal, and importance for public health strategies, is that the groups with lowest levels of knowledge, attitudes and practice also seem to be those most vulnerable to the consequences of COVID-19 infection. Older age groups are more likely to die as a result of COVID-19 and poorer groups (likely to be rural and with no education) are least likely to be able to cope with additional health care costs or loss of income due to being sick. Our findings suggest that while implementing education, awareness raising and risk communication intervention greater emphasis needs to be placed in Karnali and other rural areas targeting to isolated and vulnerable populations such as older age group, people with no education, women and widowed or singles.

Our study also showed a significant level of perceived risk among the survey population indicating that the public did not have very high expectation of easy availability of health services for them and also do not believe that government will easily control the pandemic. Participants living in rural hills, unmarried, younger age group, having higher education and working in the service sectors expressed pessimism, whilst people living in urban areas, working as labor and with little education showed more positive attitudes. Negative attitudes and high perceived risks of COVID-19 may be explained by perceptions of a slow and ineffective response by the government to the pandemic (18) and also doubts about the availability and equity of access to healthcare services (19). This is consistent with a recent study conducted in Nepal, which suggests that people tend to express negative emotions when experiencing increased anxiety and stress (9).

Despite the low knowledge and pessimistic attitudes, adherence of precautionary measures by the study population was high. People living in more developed and accessible areas, privileged caste groups, those having higher education and women reported practicing better precautionary measures.

Consistent to our finding, previous studies in other settings also showed that men compared to women, individuals with lower education and poor knowledge about COVID-19 than with higher level education and knowledge tended to practice more risk behaviors (15). Furthermore, the less precautionary measures observed among the people living in the remote areas like Karnali province could be explained by more barriers faced to adherence, such as limited water supplies, and also that adopting precautionary measures such as wearing masks and use of hand sanitizer may not be affordable.

Pertinent to this is the Nepal Demographic Health Survey finding that only 26% of households have handwashing facilities with soap and water in Karnali province (20).

Consistent to previous studies, our findings confirmed positive correlations between knowledge-practice and attitude-practice (17), however there was no correlation between knowledge-attitude. This study was implemented during the initial phase of the COVID-19 outbreak and expressed attitudes and beliefs by the study population may change as the pandemic evolves over the coming months. Higher practice scores compared to knowledge and attitude could be due to the lockdown imposed by the government that limited individual mobility and exposure to crowds. The study found that only education had significant positive correlation with knowledge, attitude and practices. It found a weak but statistically significant negative correlation between knowledge and the other demographic factors Interestingly province, ecological zone and age showed significant associations with practice after controlling confounders. These findings are in line with the results of some previous studies (21,22). Further study is recommended to reaffirm and track changing correlations between knowledge, attitude and practice, as this study was a cross-sectional survey conducted during the initial phase of pandemic outbreak.

### Strength and limitation of the study

The study sample of 618 was recruited from 26 health facilities across the country and included population groups from different strata including ecological region, caste ethnicity, and both urban and rural residents; although the proportion of urban participants (79%) was disproportionately higher than the national rate and this sampling bias may affect the generalizability of the findings.

The purposive selection of health facilities conducting fever clinics may have resulted the selection bias. Likewise, further bias may have occurred with the exclusion of people under 18 years and individuals with communication difficulties. Furthermore, the absence of visual cues on the phone might have hindered attempts to create an enabling environment for the interviews (23). We also acknowledge the possibility of social desirability bias as the data were self-reported.

## Conclusion

The findings of this study showed that the general public in Nepal have been following some precautionary practices despite their low level of knowledge and pessimistic attitude towards COVID-19 pandemic. The study demonstrated limited positive correlations between knowledge-practice and attitude-practice, but also disparities, suggesting that higher levels of knowledge and positive attitudes can result in good practices to prevent spread and remain safe during the current pandemic. There is a need to improve knowledge about COVID-19 across all parts of the population, but particularly to design and implement more targeted strategies for community-based awareness raising and risk-reduction communication intervention to enable improvements in attitudes and practices towards COVID-19 amongst the most vulnerable groups.

## Data Availability

The data used in this study will be available from the corresponding author upon reasonable request

## Declarations

The views expressed in this article are those of the authors and do not necessarily represent the views of supporting agency – Community Support Association of Nepal (COSAN).

We, the authors declare that we have no competing interest.

### Consent for publication

Not Applicable

### Availability of data and material

The datasets used for this study are available from the corresponding author upon reasonable request.

### Author’s contribution

HRD, PA and TRS conceived, designed and implemented the study in the field. RB contributed to study design and undertook data collection mobilizing enumerators. HRD analysed data and wrote the manuscript with the help of TRS and PA. AC, RB and RK provided input for finalization of manuscript and data analysis. All authors reviewed and approved the final version of the manuscript.

## Acknowledgement

The authors acknowledge the support and contribution of Community Support Association of Nepal (COSAN) and the colleagues who offered organizational facilities and staff time for this research. In addition, the authors wish to acknowledge the support provided by the hospitals and the data collectors – Shital Shrestha, BhumikaSunuwar, Nabin Basnet, Asmita KC, Aayush Shrestha, JeshikaShahi, Radhika Khadka, Dikshya Sharma, Sashi Bam, Ajay Poudel, RakshyaAdhikari, AahanaSapkota, Prativa Pandey, BibhushiBhattarai, Saleena Shrestha, AagyaDahal, AaradhanaRayamajhi, PrashabdhiShakya, NishaAdhikari, SapnaChaudhari, DeepaGhimire and Nirmala Koju. The authors are grateful to those who participated in the study and shared their views and personal experiences.

